# Requirements for the containment of COVID-19 disease outbreaks through periodic testing, isolation, and quarantine

**DOI:** 10.1101/2020.10.21.20217331

**Authors:** Ruslan I. Mukhamadiarov, Shengfeng Deng, Shannon R. Serrao, Priyanka, Lauren M. Childs, Uwe C. Täuber

## Abstract

We employ individual-based Monte Carlo computer simulations of a stochastic SEIR model variant on a two-dimensional Newman–Watts small-world network to investigate the control of epidemic outbreaks through periodic testing and isolation of infectious individuals, and subsequent quarantine of their immediate contacts. Using disease parameters informed by the COVID-19 pandemic, we investigate the effects of various crucial mitigation features on the epidemic spreading: fraction of the infectious population that is identifiable through the tests; testing frequency; time delay between testing and isolation of positively tested individuals; and the further time delay until quarantining their contacts as well as the quarantine duration. We thus determine the required ranges for these intervention parameters to yield effective control of the disease through both considerable delaying the epidemic peak and massively reducing the total number of sustained infections.

## 1. Introduction

In December 2019, a novel coronavirus, known as SARS-CoV-2, emerged in the human population and caused an on-going, widespread pandemic. COVID-19, the disease caused by SARS-CoV-2, resulted in significant losses in lives, health, and the economy. With more than 250 million cases and around five million officially confirmed deaths, at the time of writing, additional methods are needed to accurately understand and predict the spread of the disease. Mathematical models are important tools to quantify the non-linear interactions inherent to infectious disease spread, e.g., [1, 2, 3].

The dynamics of an incubation-type disease in a population can be mathematically captured by variants of the Susceptible-Exposed-Infected-Recovered (*SEIR*) model [4, 5, 6]. In the SEIR compartmental model, individuals may assume four distinct states: *S - Susceptible, E - Exposed, I - Infected*, and *R - Recovered*. Mean-field rate equations are often utilized to encode the SEIR reactions and predict how the total number of individuals in each state evolves with time. However, these coupled ordinary differential equations invoke a mass-action like factorization of higher moments into powers of compartmental population numbers, and hence neglect temporal fluctuations and/or spatial variations. Specifically, the rate equation description cannot properly account for the strong number fluctuations that drive the continuous phase transition when the system is near the epidemic threshold, nor can it account for spatially correlated clusters and spreading fronts that are induced by the disease’s propagation through nearest-contact infection [7, 8, 9, 10, 11]. Consequently, the rate equation approximation cannot capture stochastic extinction events if the disease parameters are set below or near the epidemic threshold. Furthermore, this approximation severely underestimates the ultimate prevalence of the above-threshold epidemic in the population [12, 13].

The use of different varieties of network structures has helped epidemiologists better understand the spread of infectious disease. In a commonly used network modeling framework, the complexity of the many-body interaction of society can be mapped with different structures, where each node represents an individual, and edges represent interaction among individuals [3, 14, 15]. In modern human societies, disease spreading in a confined spatial environment is more adequately described by a graph model that represents social contact interactions, and that allows for both short-range diffusive propagation as well as farther-reaching contacts along travel routes. While direct links on a lattice would enable the infection to spread from the carriers to their immediate susceptible neighbors, additional long-range connections emulate ‘express’ routes for disease transmission to spatially distant regions. For example, a detailed study of close proximity interactions in an American high school examined more than 700,000 interactions and found a network with small-world properties [16]. Realistic contact networks are rather complex, and, in fact, tend to vary from one local community to another [17]. Therefore, it is more convenient and parsimonious to work with a generic network model that is not constrained by the availability of the real-network data.

The unusual abundance of non-symptomatic infected individuals constitutes a major obstacle in containing the spread of SARS-CoV-2 [18, 19]. Such asymptomatic or pre-symptomatic individuals do not realize they are infected or transmitting and thus go about their typical daily routine, leading to additional spread of the virus. Furthermore, a scant availability of tests, delays in the process, and potential reluctance of undergoing testing and isolation procedures, may substantially undermine mitigation efforts. In practice, and especially for a highly contagious disease such as COVID-19 with a sizeable fraction of non-symptomatic infections, even with a repeated and effective testing regimen, it is impossible to capture and isolate every single infected individual. To deal with a significant fraction of unidentified infectious individuals, it is necessary to capture the direct neighborhood of confirmed disease carriers, and quarantine these individuals. That is, even if a person who had direct contact with the identified infected individual does not show any symptoms of infection, that person should still be separated from the general population. Only then can further infection propagation through unidentified (perhaps asymptomatic) disease carriers be controlled effectively.

Our goal here is to investigate the robustness of the intervention schemes for periodic testing and isolation regimen of both infected individuals and their contacts. While this central idea of repeatedly screening the population, isolating positively tested individuals, and quarantining their immediate contacts, irrespective of their infection status, is straightforward and long-established, its efficacy depends on the details of the spreading environment and on the ability to determine who is infectious in a timely manner [20, 21, 22]. Since outbreaks usually take place within some smaller-scale communities, in this work, we consider an isolated population of the size of a small town or university / college that follows social distancing guidelines and is subject to a mask mandate, i.e., a pool of individuals of the size of a few tens of thousand individuals with diminished number of contacts and thus reduced chances of transmitting the infection. As for the delays in testing and quarantining, for the current epidemic, they occur mainly due to the following three reasons: (1) there is a time delay before receiving test results; (2) there is an (unavoidable) time lapse between the identification of infectious individuals and their placement in isolation; and (3) subsequently it takes a finite time until all infectious persons’ contacts are identified and quarantined. To properly incorporate the impact of these distinct delays on disease transmission in a heterogeneous network, we study the spread of SARS-CoV-2 using a fully stochastic and properly spatially extended representation of the SEIR model. In order to demonstrate how the subtle details of a chosen spatial (or network) setting affect our qualitative observations, we run our individual-based Monte Carlo simulations on three different variants of small-world networks.

We find that for all three distinct small-world networks that we have considered, the fraction of identified infectious individuals and the testing period both play decisive roles in containing the infection outbreak. In contrast, within reasonable bounds, the delays in testing and quarantining, as well as the quarantine duration, affect the course of the outbreak in a much more limited manner. In fact, some of the latter mitigation parameters hardly impact the disease dynamics at all while the others do so more if the individuals are allowed to move across the network, and vice versa if the network is static. Therefore, in prioritizing resources to achieve an optimized testing / isolation / quarantine strategy, primary emphasis should be placed on frequent and comprehensive testing, less on reducing minor delays in reporting test results and effective contact tracing. Finally, after varying both the fraction of identified infectious individuals and the testing period simultaneously, we located the crossover region which separates the presence and absence of a major outbreak.

## 2. Model description

### 2.1. Modified stochastic SEIR model on a Newman–Watts small-world network

In the underlying SEIR formulation, the disease spread is represented by three characteristic reactions that we consider as independent stochastic processes: *S* + *I* → *E* + *I* with the reaction rate *r, E* → *I* with the reaction rate *b* (given by the inverse incubation period), *I* → *R* with reaction rate *a* (given by the inverse recovery period). For comparison, the associated dynamical mean-field rate equations are listed in the appendix, eqs. (A.1)–(A.5). The basic reproduction number for the system is proportional to the ratio of the infection to the recovery reaction rates ℛ_0_ ∝ *r/a*, with a proportionality constant that is given by the mean number of contacts of each individual [23]. To capture potential limitations for effective testing and isolation protocols in our mathematical model, we replace the infectious state in the SEIR model with two distinct configurations that we term *identified infectious* (*I*) and *unidentified infectious* states, respectively. Here, ‘identification’ is to be understood as an umbrella term that encapsulates the overall capability of the testing procedure to identify and subsequently isolate infected individuals. That is, if the entire population is tested, then only the fraction of identified infectious individuals will be detected and quarantined, i.e., temporarily removed from the system, which has the effect of disrupting their future infection chain with other susceptible individuals. Likewise, the *A* state can also be viewed as a surrogate for those infectious individuals that are missed out by a standard testing procedure. In this respect, any unidentified infectious individuals will continue to spread the disease until they ultimately recover from it. The schematics of this modified SEIR model variant is displayed in Fig. 1. The reactions *E* → *I* and *E* → *A* may occur with the respective rates *f b* and (1 − *f*)*b*, where *f* denotes the fraction of identified infectious individuals, an important independent parameter in our study. Note that we assume identical infection and recovery rates for individuals in both the identified or unidentified states.

**Figure 1.**
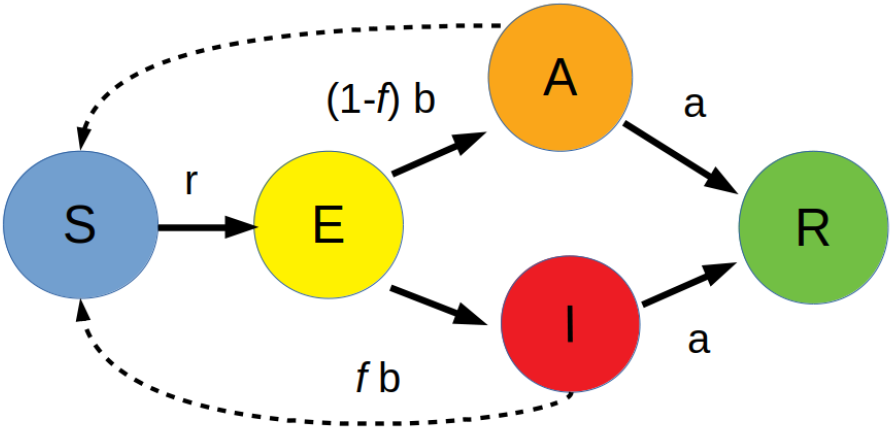
Flow diagram for the modified SEIR model. The arrows between different states represent the stochastic reactions with the appropriate reaction rates. Our modification entails replacing the infectious state with distinct *identified infectious* and *unidentified infectious* states. The fraction *f* of identified infectious individuals constitutes a crucial adjustable parameter in the model. The dashed arrows indicate that interaction of a susceptible individual with either an identified or unidentified infectious individuals is necessary for infection.

While the infection dynamics may depend on the structure of the underlying spatial setting, in this work we wish to demonstrate that our intervention scheme, which we introduce in the next subsection, is in fact robust against fine variations in details of the employed network. To approach this task, we utilize dynamic and static variants of Newman–Watts small-world network [11] on different underlying two-dimensional lattices as a suitable spatial framework for our individual-based Monte Carlo simulations of the COVID-19 epidemic propagation [24, 25, 13]. Below we list all three variations of the two-dimensional Newman–Watts small-world network that we consider in this work:

i. The first variation consists of a regular King’s graph with *L* sites in each dimension, and additional long-distance links, c.f. Fig. 2a. We choose a King’s graph to achieve a more realistic number of contacts for each individual during each simulation cycle. Thus, it comprises 2(2*L*^2^ − 1) short-range links (where the additional 6*L* − 4 links stem from the periodic boundaries) that connect the nearest-neighbor lattice vertices, and 4*ϕL*^2^ additional long-distance links between randomly selected lattice sites. Here, *ϕ* denotes a prescribed probability for a long-range link to form, and the average connectivity of the network hence is ⟨*k*⟩ = 8(1 + *ϕ*). In order to minimize finite-size effects, we set the lattice size in each dimension to be *L* = 1000, and employ periodic boundary conditions; i.e., close the square lattice to a two-dimensional torus. Each of the *L*^2^ lattice sites may be occupied by at most a single individual, labelled by the distinct states *S, E, I, A*, or *R*. For this network variant we allow all individuals to move around the lattice by letting them hop with the rate *d >* 0 to other empty connected lattice sites. Therefore, we henceforth refer to this spatial network organization as a dynamic small-world network on a King’s graph or simply as a *dynamic King’s graph* (DK) setup.

**Figure 2.**
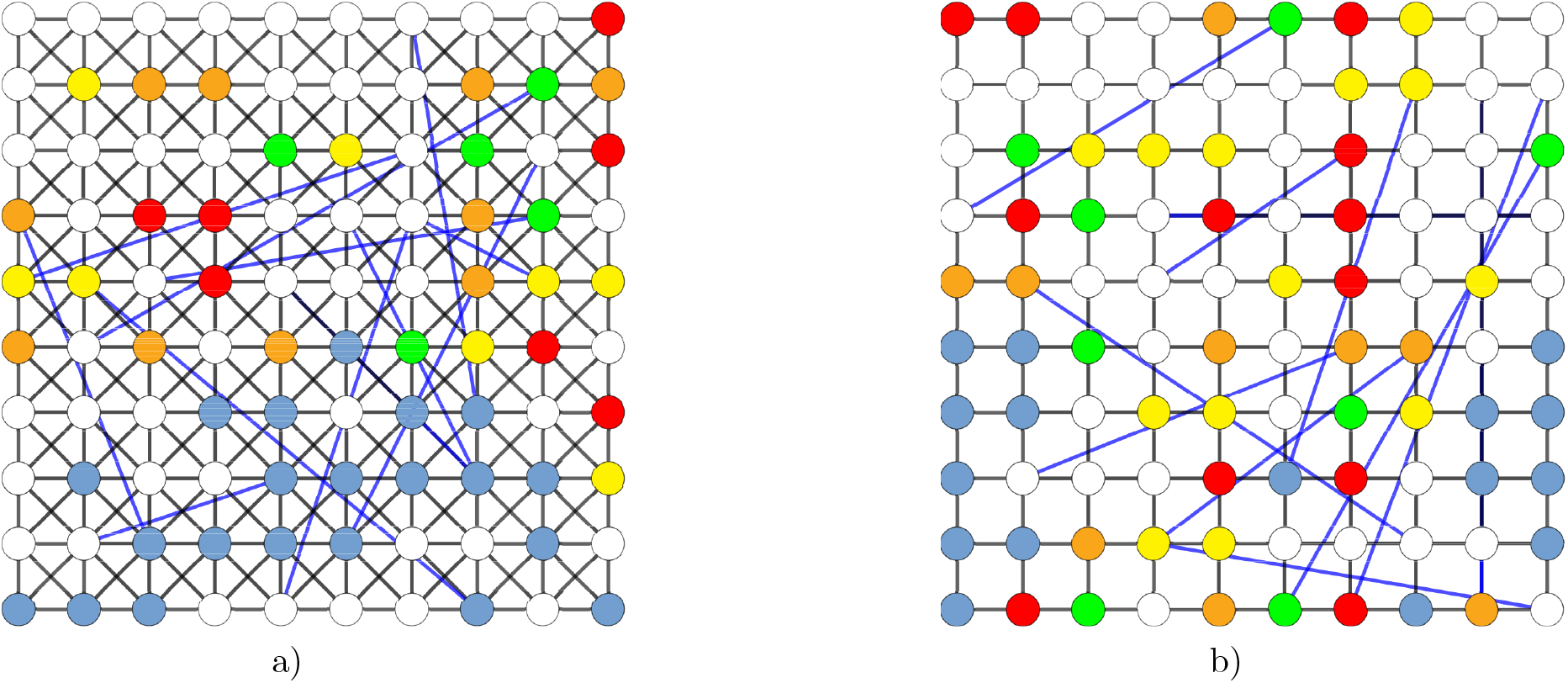
Schematic construction of a Newman–Watts small-world network in two spatial dimensions (with periodic boundary conditions, i.e., on a torus), obtained from (a) a regular King’s graph or (b) from a regular lattice, through adding long-distance connections (blue) between randomly selected lattice sites. The graph displays a modified SEIR model configuration snapshot with sites residing in empty (white), susceptible (light blue), exposed/incubative (yellow), identified infectious (red), unidentified infectious (orange), or recovered (green) states (color coding as in Fig. 1).
ii. The second network variant we consider is a special case of the first one *without* hopping, i.e. *d* = 0, which we refer to as static small-world network on a King’s graph or simply as a *static King’s graph* (SK) setup.
iii. We also consider a completely different spatial organization by generating the Newman–Watts small-world network on a regular two-dimensional lattice, c.f. Fig. 2b, for which the average connectivity of the network reads ⟨*k*⟩ = 4(1 + *ϕ*). As for the dynamic King’s graph, here we also allow all individuals to move around the lattice by letting them hop with the rate *d >* 0 to other empty connected lattice site. Thus, we refer to this network as *dynamic square lattice* (DS) setup.

In order to compare the results between these three networks, we set the basic reproduction number for all three setups to ℛ_0_ ≈ 2.5, which is achieved by varying the total population density *ρ*_pop_.

Once the combined *N* individuals of *S* and *I* species have been randomly distributed over the small-world network with fixed initial density, the simulation proceeds with parallel sequential updates. A selected individual is allowed to react and/or to move around if the network is dynamic, subject to the set of possible individual reactions prescribed by our stochastic agent-based modified SEIR model. (The detailed stochastic simulation algorithm is described in the Appendix.) The control parameters that regulate the speed of the unmitigated infection spread are: the initial population density in each state, the values of the reaction rates depicted in Fig. 1, and the average connectivity ⟨*k*⟩ of the small-world network. On the lattice, upon encounter of a susceptible *S* with an infectious individual *I* or *A*, one of the binary reactions *S* + *I* → *E* + *I* and *S* + *A* → *E* + *A* transforms the susceptible into the exposed state *E* with the infection rate *r*. Hence this choice defines the intrinsic simulation time scale.

To model the COVID-19 epidemic on small-world networks, we set the incubation rate to *b* = (1*/*6) days^*−*1^, and the recovery reaction rate as *a* = (1*/*6.5) days^*−*1^, where 1*/b* and 1*/a* represent the mean incubation and recovery periods, respectively, as reported for this disease [22, 26, 27, 28]. As these rates reflect biological characteristics of COVID-19, we fixed them and chose other parameters such that the population undergoes epidemic outbreaks for all three different choices of the underlying network, i.e., all three systems are set above the epidemic threshold when there is no intervention measure imposed, with an effective basic reproduction number tuned to ℛ_0_ ≈ 2.5, again informed by estimates for COVID-19 [22, 29, 30, 31, 32]. To facilitate comparison across network settings, we fixed the probability for the formation of long-range links to *ϕ* = 0.6, the infection rate *r* = 0.6, and the probability of moving *d* set to 1 for dynamic networks and 0 for static setups. The general rule of thumb that we followed when picking a value for each of these parameters was to stay away from the limiting cases. For instance, the lower bound on the system’s density *ρ*_*pop*_ is set by the epidemic threshold, while the upper bound is chosen in such a way that one could still look at the effects of agent hopping. As for the probability for the formation of long-range links *ϕ*, we select its value from the range where both total number of recovered individuals and the peak of the infectious curve seems to saturate (see Fig. 3). Finally, the values for the infection rate *r* and the hopping rate *d* are chosen in such way that the peak and the width of the resulting infectious curve fit well to the mean-field infectious curve obtained by setting the recovery rate *γ* = *a* = (1*/*6.5) days^*−*1^, the incubation rate *α* = *b* = (1*/*6) days^*−*1^, and the infectious rate *β* = ℛ_0_*/γ*, where ℛ_0_ ≈ 2.5.

**Figure 3.**
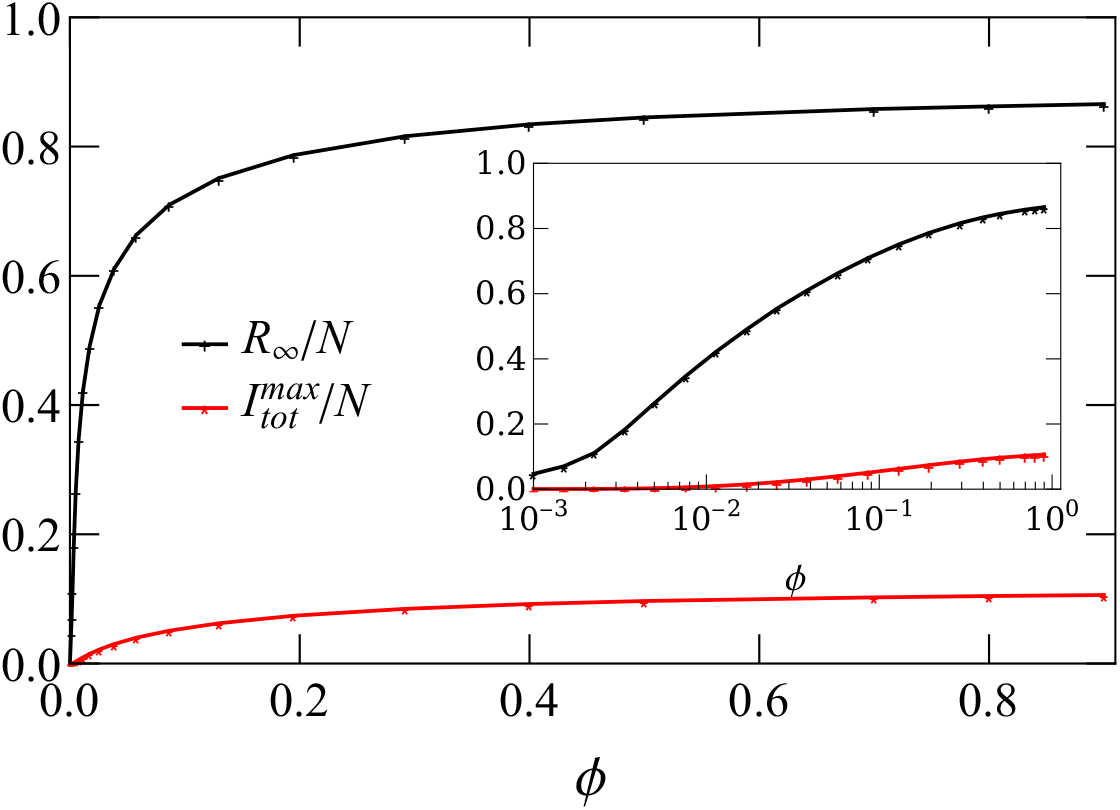
The eventual recovered population and the peak of the infected population for the unmitigated dynamic King’s graph both saturate quickly as the probability for the formation of long-range links increases. Without loss of generality, we choose *ϕ* = 0.6 throughout this paper. The inset shows the same data with a logarithmic scale on the *ϕ* axis.

Thus, to achieve ℛ_0_ ≈ 2.5, we varied the population density *ρ*_*pop*_. For the dynamic small-world network on a King’s graph, this entails a population density of *ρ*_pop_ = 0.59 of the total number *L*^2^ of lattice sites (as *L* = 1000, here, *N* = 590, 000). Thus, the average number of contacts for each individual per day is estimated to ⟨*k*⟩*ρ*_pop_ ≈ 7.6, which is in agreement with the empirical estimation of 5 to 20 contacts per day [33, 34]. For the static setup with *d* = 0, in order to maintain ℛ_0_ ≈ 2.5 for the unmitigated runs, we have to increase the number of long-range connections by setting *ρ*_pop_ = 0.73, which also raises the average number of contacts to 9.3. Since for the dynamic small-world network on a regular lattice the number of short-range links is lower than in the King’s graph, we achieve ℛ_0_ ≈ 2.5 at a higher value of the population density *ρ*_pop_ = 0.62 than for the dynamic King’s graph, amounting to a total population of *N* = 620, 000. With the underlying square lattice now containing 2*L*^2^ short-range links in presence of the periodic boundaries and 2*ϕL*^2^ long-range links, the network has an average connectivity ⟨*k*⟩ = 4(1 + *ϕ*), giving rise to an average number of contacts ⟨*k*⟩*ρ*_pop_ ≈ 3.97 per day. We summarize the parameter values for the different spatial setups in Table 1.

**Table 1.** Simulation parameters for stochastic SEIR model for different spatial setups (see description in the main text). Each parameter set was chosen in such way that the basic reproduction number would match the reported value for COVID-19 ℛ_0_ ≈ 2.5. The same lattice size *L* = 1000 was used for all three frameworks.

| Setup | $\rho_{\text{pop}}$ | $\phi$ | $r$ | $a$ | $b$ | $d$ |
| --- | --- | --- | --- | --- | --- | --- |
| Dynamic King's graph | 0.59 | 0.6 | 0.6 | 1/6.5 | 1/6 | 1.0 |
| Static King's graph | 0.73 | 0.6 | 0.6 | 1/6.5 | 1/6 | 0.0 |
| Dynamic square lattice | 0.62 | 0.6 | 0.6 | 1/6.5 | 1/6 | 1.0 |

### 2.2. Mitigation strategy

For the current epidemic, delays occur due to the period before receiving test results, as well as the (unavoidable) time lapse between the identification of infectious individuals and their placement in isolation, and the subsequent time until all infectious persons’ contacts are quarantined. As is illustrated in Fig. 4, there are several parameters that can be varied in this scenario: the testing period (TP); the delay between administering the test and isolation of the infected identified individual (DT); the additional delay between isolating positively tested individuals and quarantining their direct contacts (DQ); and the duration of the quarantine (Q).

**Figure 4.**
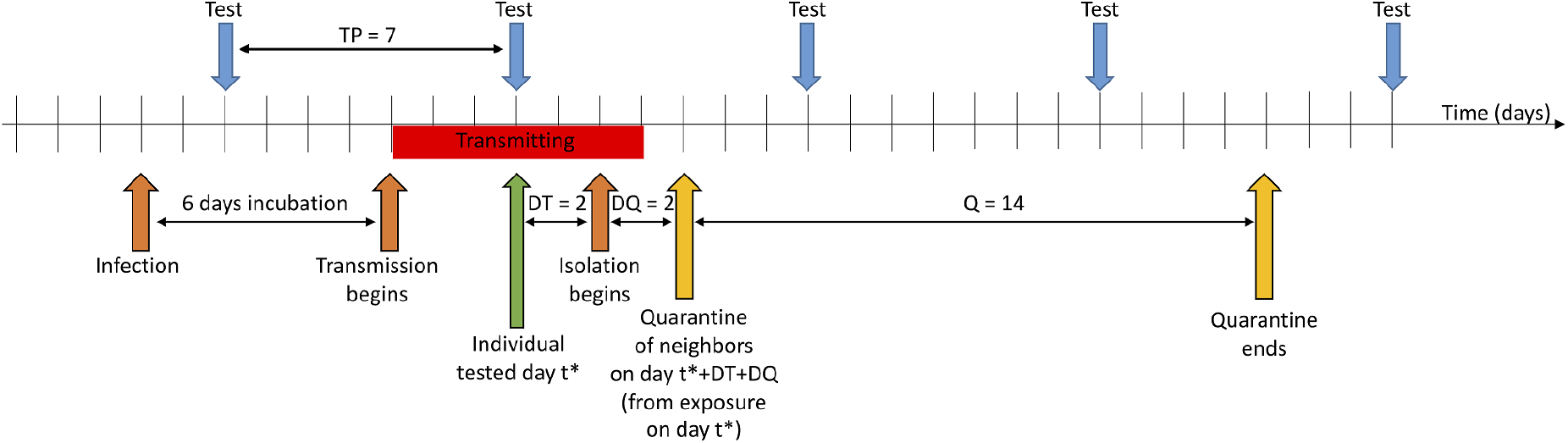
Schematic for the proposed mitigation strategy. Vertical arrows indicate various external actions on the population. The frequency of these control measures and the time delays between the different interventions represent the crucial control parameters for containing the infection. In this schematic, we show the standard value of parameters found in Table 2. Prior to the time indicated by the first orange arrow, the individual is susceptible, *S*. Upon infection, the individual enters the exposed state, *E*, where they are infected but not infectious. After six days, the individual enters the identified infectious classes, *I*. Following an additional six days, the individual recovers to state *R* and is no longer infectious. This sketch refers to one representative individual; the length of the incubation and infectious periods are varied in this study.

In the present study, we thus consider the following mitigation protocol: To be able to capture identified infectious individuals, the population needs to be regularly tested with some prescribed period TP. Once the periodic testing measure in the community begins at some time instant *t*^*\**^ (subsequently repeated according to the description provided in the next paragraph), the entire population, irrespective of the state each individual happens to be in at the time, is considered screened (i.e., full information is available in the model). Only the identified infectious individuals, however, are registered. These individuals are placed in isolation after some fixed delay time DT. Contact tracing is implemented in our model through registering the information about each identified individual’s neighbors along any of the links in the small-world network, at the time *t*^*\**^ of the testing. At the later time *t*^*\**^ + DT + DQ, these nearest neighbors will be placed in quarantine, where they cannot interact with any of their neighbors, and they cannot move (if *d >* 0). Subsequently, both the originally identified infectious individuals and their nearest neighbors will be released from quarantine after Q days from the moment they have been isolated. Note that these individuals may change their state from *E* to *I/A* and *A* or *I* to *R* with the corresponding fixed rates while in quarantine or isolation. For example, infectious individuals in isolation recover with rate *a*.

In order to clearly demonstrate the efficacy of this mitigation strategy, it is necessary to set the initial conditions of our model to admit sizeable outbreaks to occur for all three small-world network variants that we consider here. To this end, we take the initial number of the infection centers in the system to be *I*_0_ = 10, and we start to test and isolate the individuals 10 days after these infection nuclei were planted in the population. Furthermore, we set the basic model parameters that describe the testing protocol to certain realistic default values, also listed in Table 2; namely *f* = 50%: one half of the infectious population is identifiable through the testing, a ratio that incorporates both test availability and reliability; TP = 7 days: periodic testing campaigns are carried out weekly; DT = 2 days: it takes two days for the tests to be evaluated and to arrange for the infectious individuals to be put into isolation; DQ = 2 days: subsequent contact tracing consumes another two days until the immediate contacts (through either short- or long-range links in the small-world network) are quarantined; and Q = 14 days: isolated and quarantined individuals are kept immobile and disconnected from the remainder of the population for two weeks.

**Table 2.** Control parameters varied during mitigation. Figure 4 provides an example of how these variables operate and affect the intervention scheme.

| Parameter | Description | Standard Value | Range |
| --- | --- | --- | --- |
| $f$ | identified infectious fraction | 50% | 10 – 95 % |
| TP | testing period | 7 days | 1, 2, 5, 7, 10 days |
| DT | delay to testing | 2 days | 1, 2, 5, 7, 10 days |
| DQ | delay to quarantine | 2 days | 1, 2, 5, 7, 10 days |
| Q | quarantine length | 14 days | 5, 7, 10, 14, 21 days |

## 3. Results

In this section we evaluate the effectiveness of our mitigation strategy for the three different small-world networks described above. Running Monte Carlo simulations for these stochastic SEIR model variants, we find that our choice of the spatial setting does not significantly change the course of unmitigated disease spread provided we select the simulation parameters such that all different setups lead to the same basic reproduction number ℛ_0_. However, we show in Fig. 5 that depending on the setup, the disease spread may be suppressed quite substantially when the mitigation measures are introduced. As is displayed in the figure, the disease spread on the small-world network on a King’s graph without diffusion, i.e., on the static setup, is suppressed the most by our contact tracing and isolation control strategy. That makes sense since by setting the hopping rate to zero (*d* = 0) we effectively prohibit the individuals to change their neighbors, or in other words, we fix the existing set of connections between individuals. Since for that system the set of neighbors does not change, isolating the infectious individuals and its neighbors amounts to effectively removing the disease spreading center. At the same time, when we set *d >* 0 we allow individuals to rewire their connections through their diffusion across the network, which permits the infectious individuals to come into contact with many more susceptible individuals, accelerating disease spreading. We note that due to the periodic testing protocol and the subsequent imposed mitigation measures, both the fraction of quarantined and the fraction of isolated individuals oscillate over time. To obtain statistically meaningful results, all measured quantities were averaged over 100 runs in the results reported below. The ensuing statistical error bars that we have computed from our data are in fact smaller than the size of the plot markers and hence cannot be clearly discerned in the graphs.

**Figure 5.**
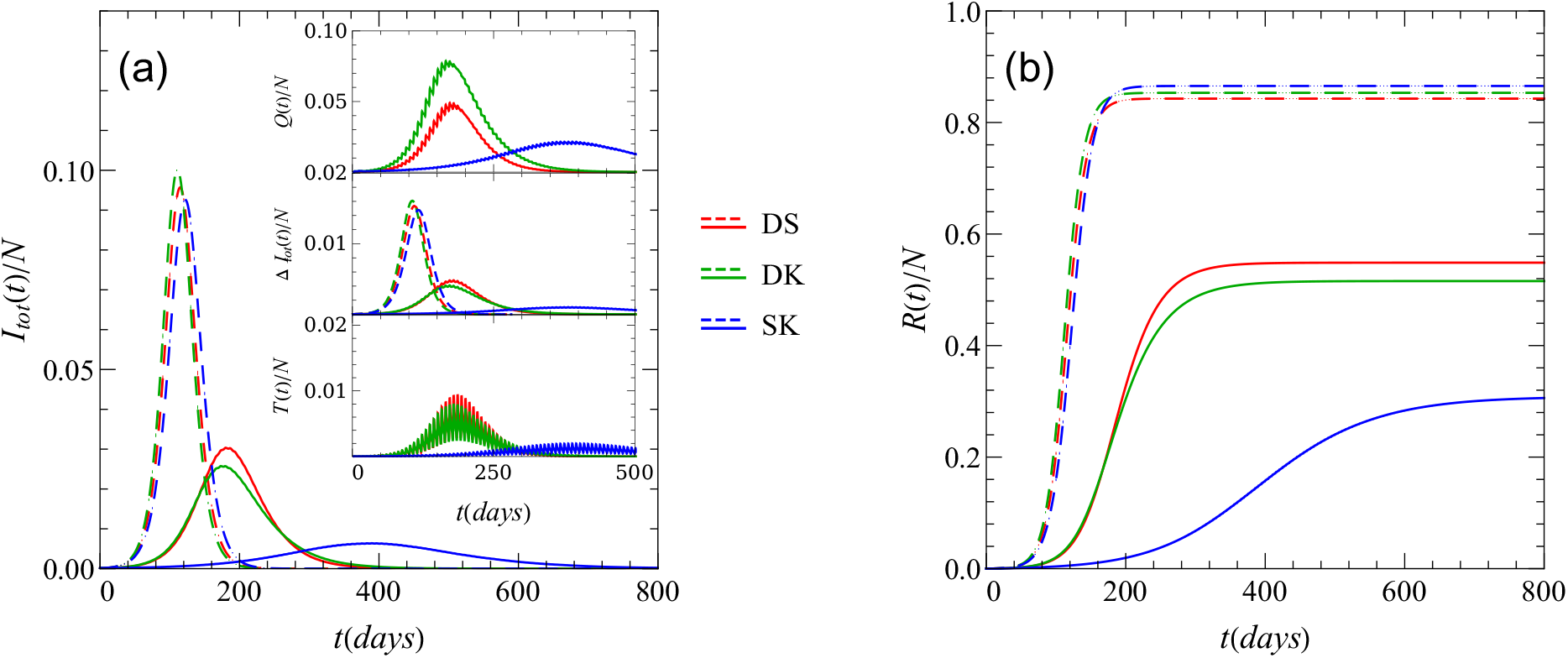
Mitigated and unmitigated time evolution of (a) the total fraction of infectious (symptomatic and asymptomatic combined) and (b) recovered individuals for three different choices of the underlying network. The acronym DS stands for dynamic square lattice, DK for dynamic King’s graph, and SK for static King’s graph. The insets additionally display the fractions: *T* (*t*)*/N* of isolated individuals, Δ*I*_*tot*_(*t*)*/N* of daily new infections, and *Q*(*t*)*/N* of quarantined individuals. For all mitigated curves we used the standard values of the mitigation parameters that are shown in Table 2. The dashed curves correspond to unmitigated dynamics.

To estimate the impact of each control parameter on the course of the disease dynamics, we fixed all parameters to their standard values (see Table 2), and then varied each one of them separately, measuring the peak of the infectious curve 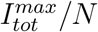 and the fraction of total recovered population *R*_*∞*_*/N*. First, we varied the fraction of identified infections individuals: as is evident in Fig. 6, an increase in *f* drastically reduces both fractions 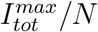 and *R*_*∞*_*/N*. For the static setup, our results show that the infection spread is entirely quelled for *f* ≥ 0.75 (and other parameters set at their standard values). Similarly, as shown in Fig. 7, decreasing the testing period TP leads to a significant drop in both 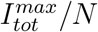 and *R*_*∞*_*/N*, and even halts the disease spread completely for the static case when TP≤ 2 days. In contrast, variation of the testing delay parameter DT does not affect the infection dynamics at all for the static case and only influences the outbreak in networks that allow diffusive motility, c.f. Fig. 8. As for the quarantine duration Q and the delay to quarantine DQ, the dynamic network settings (*d >* 0) appear to be quite insensitive to variations in the parameters Q and DQ, see Figs. 9 and 10. This observation suggests that continuous rewiring of existing connections in a network renders the isolation of the neighbors of the infectious individuals ineffective.

**Figure 6.**
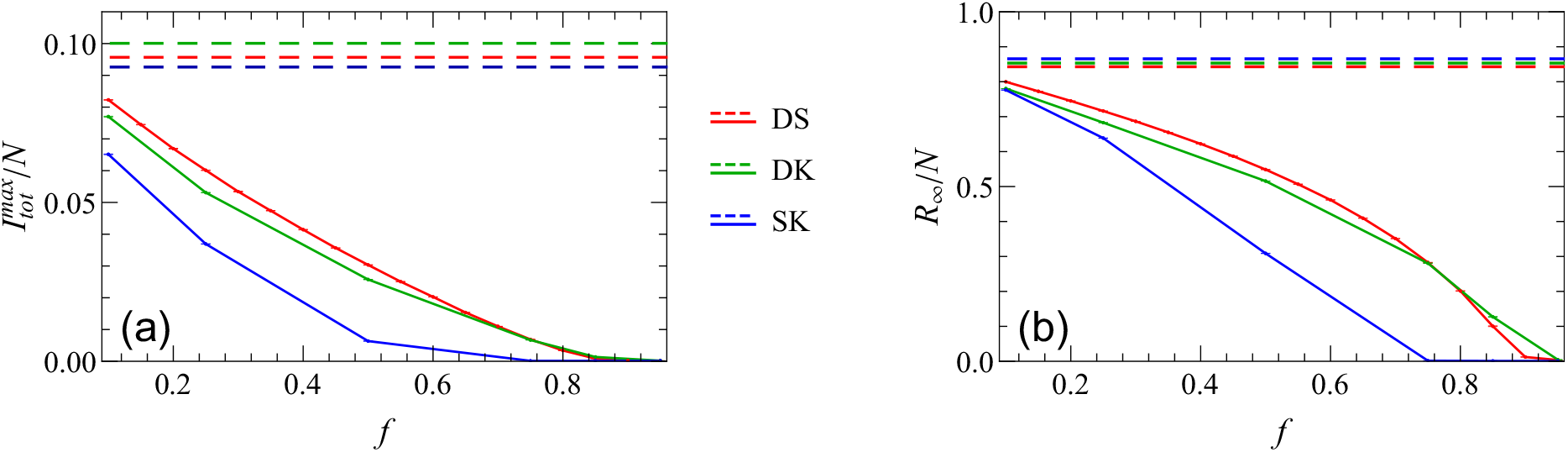
Change of (a) the peak of the infectious curve 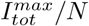 and (b) the fraction of total recovered population *R*_*∞*_*/N* with the fraction of identified infectious individuals *f* for three different setups. The other mitigation parameters are set to their default values (TP = 7, DT = 2, DQ = 2, Q = 14). The dashed lines represent unmitigated scenarios.

**Figure 7.**
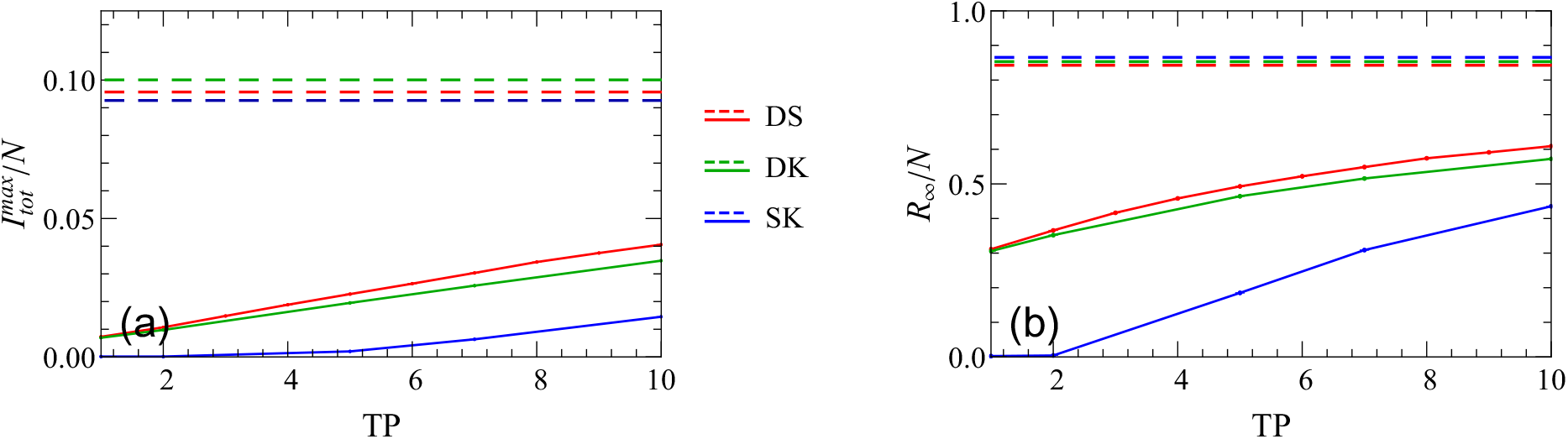
Change of (a) the peak of the infectious curve 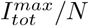 and (b) the fraction of total recovered population *R*_*∞*_*/N* with the testing period TP for different setups. The other mitigation parameters are set to their default values (*f* = 0.5, DT = 2, DQ = 2, Q = 14). The dashed lines represent unmitigated scenarios.

**Figure 8.**
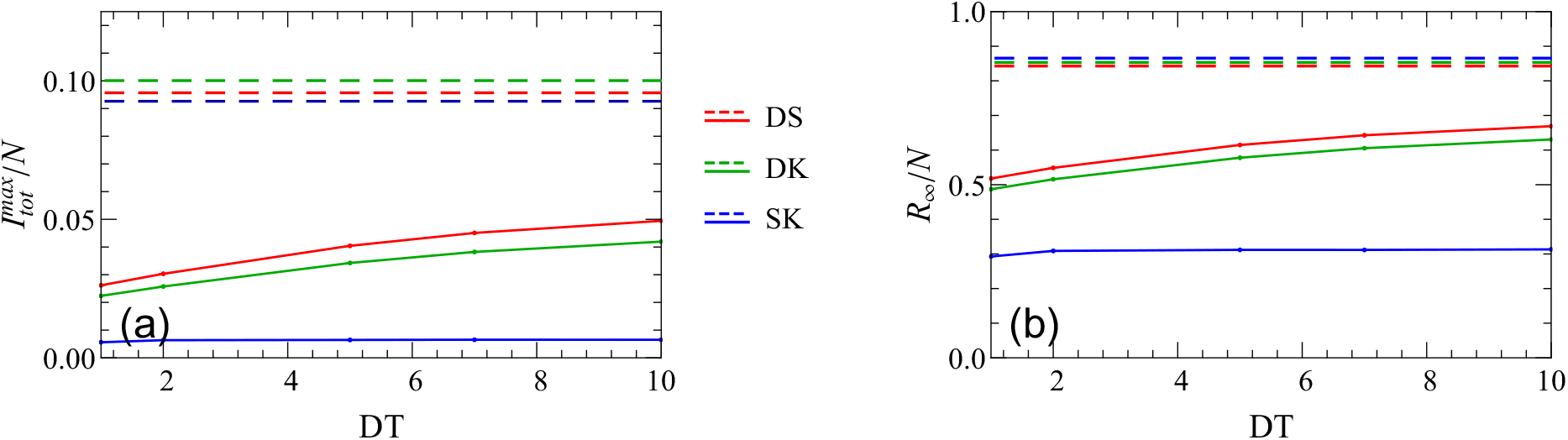
Change of (a) the peak of the infectious curve 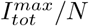 and (b) the fraction of total recovered population *R*_*∞*_*/N* with the testing delay DT for different setups. The other mitigation parameters are set to their default values (*f* = 0.5, TP = 7, DQ = 2, Q = 14). The dashed lines represent unmitigated scenarios.

**Figure 9.**
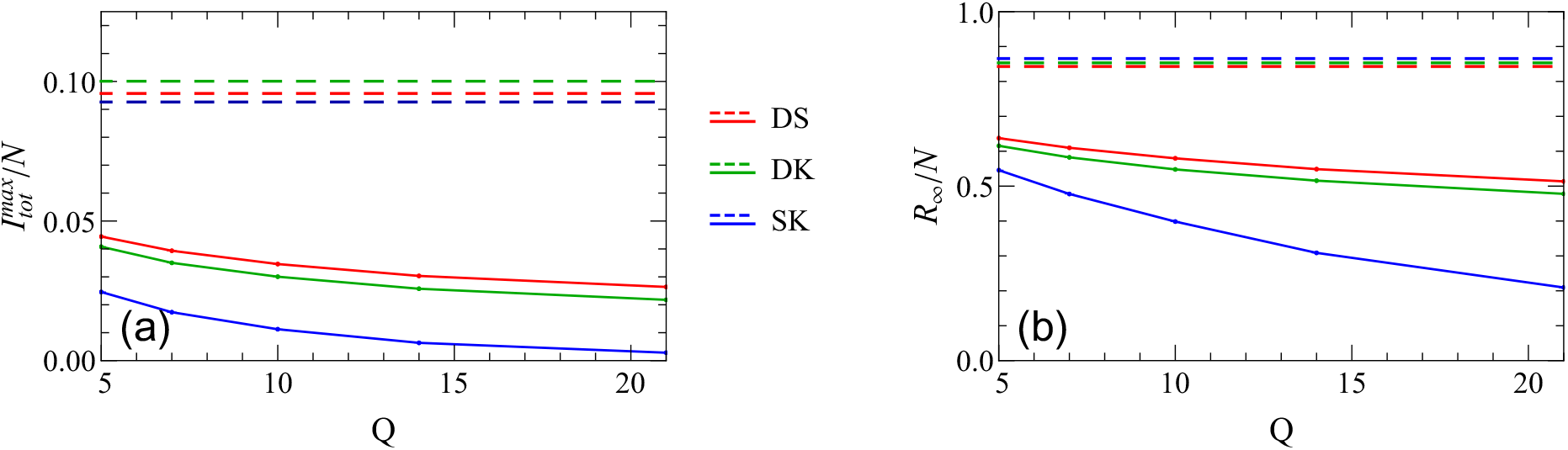
Change of (a) the peak of the infectious curve 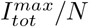 and (b) the fraction of total recovered population *R*_*∞*_*/N* with the quarantine duration for different setups. The other mitigation parameters are set to their default values (*f* = 0.5, TP = 7, DT = 2, DQ = 2). The dashed lines represent unmitigated scenarios.

**Figure 10.**
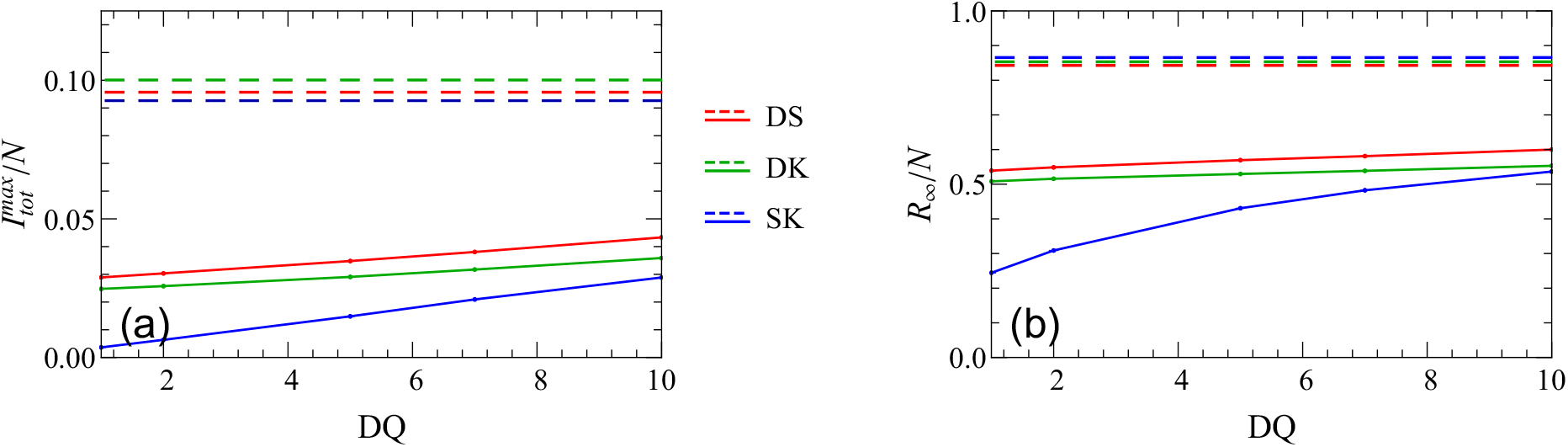
Change of (a) the peak of the infectious curve 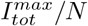 and (b) the fraction of total recovered population *R*_*∞*_*/N* with the delay to quarantine for different setups. The other mitigation parameters are set to their default values (*f* = 0.5, TP = 7, DT = 2, Q = 14). The dashed lines represent unmitigated scenarios.

Thus far we have demonstrated that according to our mitigation scheme, varying the fraction of identified infectious individuals *f* and the testing period TP influences the course of the disease dynamics and its final outcome considerably. Therefore, to summarize our quantitative parameter study data, we plotted three sets of heat map plots in Fig. 11, one for each setup, where we have shown how the peak in the curve of the fraction of infected individuals, the fraction of the total number of recovered individuals at the end of the outbreak, and the time at which the infection peaks in the population, are changing with *f* and TP, respectively. These graphical representations indicate that for low values of *f* and high values of TP, the outbreak remains essentially unmitigated for all three network settings that we have considered here.

**Figure 11.**
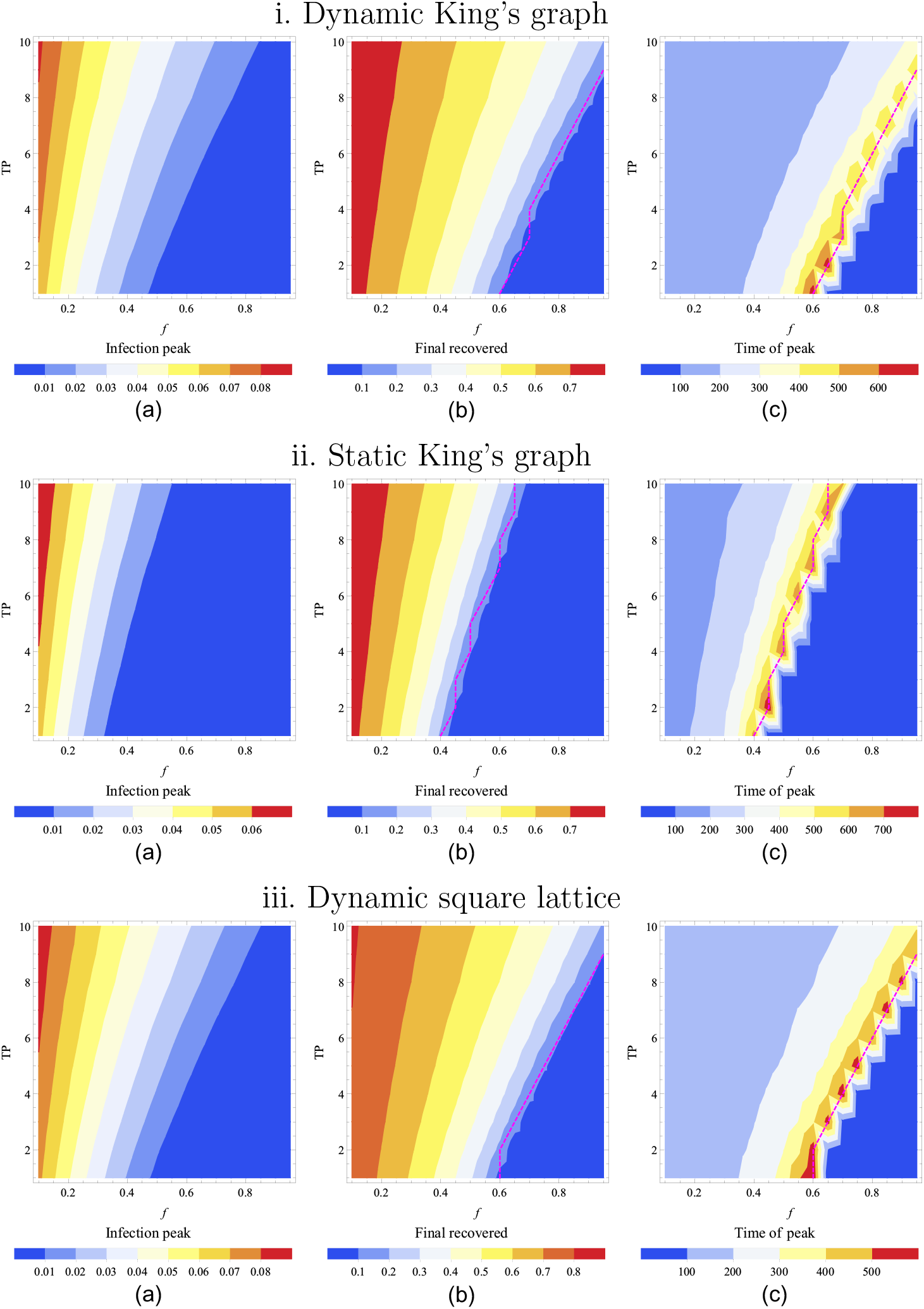
Heat map representations of pertinent outcomes for the three setups. The heat map colors respectively represent (a) the peak in the curve of the fraction of infected individuals, (b) the fraction of the total number of recovered individuals at the end of the outbreak, and (c) the time (in days) at which the infection peaks in the population. The fraction of identified infectious individuals (*f*) is varied along the horizontal axis and the testing period (TP, in days) is varied along the vertical axis. In all mitigated curves, quarantine duration of nearest neighbors, and delays between test and isolation and between isolating positively tested individuals and quarantining their direct contacts are held at their standard values (see Table 2). The rows show the scenarios for the dynamic King’s graph, the static King’s graph and the dynamic square lattice, respectively. The dashed lines indicate the position of the threshold ℛ_*E*_ = 1.

Remarkably, the heat maps in Fig. 11 appear to reveal a crossover region which marks the location of the epidemic threshold (dashed line in Fig. 11), where the value of the mitigated reproduction number ℛ_*E*_ = 1 [35, 36]. This threshold evidently coincides with the maximum infection peak time shown in Fig. 11 (c). Hence, a drastic increase in the time it takes to reach the infectious peak indicates that the system passes through a region in parameter space where the dynamics is slowing down substantially. Moreover, comparing the second and the third columns in Fig. 11, we notice that the sharp drop in the final number of recovered individuals occurs in the parameter region where the peak time displays its maximum values. These two observations lead us to the conclusion that the maximum values of the time it takes to reach the infectious peaks marks the parameter region where the system experiences a qualitative change from a large outbreak to a decaying outbreak state.

## 4. Discussion

We note that, for example, recent work by Kretzschmar et al. [37] demonstrated that a combination of non-pharmaceutical interventions, i.e., both physical distancing and contact tracing can reduce the effective reproduction number below the epidemic threshold. Varying the testing and tracing coverage as well as testing and tracing delays, the authors showed that testing delays can significantly increase the potential for onward transmission. With no delay in testing, nearly 80% of transmission is averted while only 5% is averted with a five day delay. They conclude that an advanced contact tracing method via mobile app, which minimizes testing and tracing time delays, is sufficient to quell the infection spread when the testing coverage exceeds 60%. Indeed, it has been claimed that more restrictive, widespread control measures can be substantially alleviated if testing and contact tracing programs are implemented effectively. Similar to our finding, Grantz et al. [38] determined that test effectiveness constitutes a crucial parameter for containing infectious spreading. According to their simulation results, at least 60% of the infectious population must be captured with testing (assuming perfect quarantine) for spread of the infection to be brought below the epidemic threshold. With more than 50% of infectious individuals identified, we observe significant reductions in the peak of infections and the total number that becomes infected during the epidemic (Fig. 6). In contrast, our results require a much larger fraction of infectious individuals to be identified before the systems falls below the epidemic threshold.

Our work assumes perfect adherence to quarantine, i.e., all neighbors of identified infectious individuals are unable to interact with others for the defined quarantine period. Importantly, even for perfect adherence to quarantine not all transmissions are averted as some will occur prior to quarantine and isolation. Similar results were reported by Quilty et al. [39], who determined the amount of transmission potential during quarantine or isolation, i.e., the integral of the infectivity curve over time spent in quarantine and post-quarantine isolation, weighted by compliance. They extensively explored the effects of quarantine adherence and suggested that with lower adherence to quarantine, as has been reported in the UK, only a small fraction of potential transmission is averted [40]. Both our work and that of Quilty et al. [39] note that quarantine periods of ten days rather than fourteen may be nearly as effective at averting transmission assuming high compliance.

All three studies, Refs. [37, 38, 39], utilize variations of the well-known branching process for stochastic modeling of the COVID-19 spread. However, similarly to other SEIR-based analytic studies [41, 42, 43], these branching processes were not implemented in a spatial or network setting. Therefore, these results do not account for potentially decisive spatial correlations that emerge in realistic disease spreading. It is therefore natural and relevant to ask next whether the infectious disease dynamics becomes qualitatively altered if considered in spatially extended models. This question was explored in the recent work by [13], where authors performed individual-based numerical simulations of stochastic Susceptible-Infectious-Recovered (SIR) model variants on four distinct spatially organized lattice and network architectures. They found that highly connected networks closely follow mean-field SIR rate equations, while the disease spread on a lattice and small-world network revealed marked correlation effects. A distinct investigation confirms that the dynamical behavior of infection spreading on networks with the same connectivity distribution could still differ, depending on the networks’ subtle construction details such as degree of clustering [44]. Following their results for numerical simulations of stochastic Susceptible-Infectious-Recovered-Susceptible (SIRS) model on a networks with exponential connectivity distribution, some quantities such as the mean number of infected individuals at stochastic equilibrium change with the fine details of the network structure, while others like the basic reproduction ratio ℛ_0_ appear to be completely determined by the network’s mean connectivity and the connectivity distribution.

Furthermore, we emphasize that our results in this work focus on the effectiveness of a periodic testing and quarantine regimen to contain the epidemic without implementing a lockdown. Relaxing the stringency of social distancing and contact-reducing interventions typically leads to a resurgence of the epidemic outbreak [45, 46, 47]. Recurrent second, third (and fourth) waves have occurred in many countries of our highly interconnected globe over the past months; therefore, a continuous adjustment of the efficiency of combined non-pharmaceutical interventions (testing with isolation and quarantine; lockdown) is required, depending on the population transfer between different places.

## 5. Conclusion

From our individual-based numerical simulations of infectious disease spreading on realistically motivated various small-world network architectures under partial control through testing and isolation schemes and quarantine of nearest neighbors, we draw the conclusion that, regardless of the simulation setup specifics and choice of network graph structure, a targeted improvement of mitigation protocols through systematic reduction of the testing period and maximization of the fraction of identified infectious individuals constitutes the most effective way of mitigating epidemic outbreaks. We demonstrated in our heatmap plots that deficiencies in test reliability and accuracy can be compensated by increased testing frequency, thus pushing the system towards a non-endemic state.

At the same time, delays in effecting isolation affect the disease spread only when it is simulated on a dynamic network, while delay in quarantine, and shortening the quarantine period itself, appear to have an impact only on static networks. Nevertheless, our simulations demonstrate that these three delay parameters (DT, Q, DQ) are not as sensitive in reducing the infection peak and the cumulative number of infected individuals in the population; these variables should hence be considered with lower priority for efforts to optimize testing mitigation protocols.

## Data Availability

Data is found in the publicly available git hub repository: https://github.com/shannonserrao/COVID_SEIR

## Acknowledgments

Research was sponsored by the U.S. Army Research Office and was accomplished under Grant Number W911NF-17-1-0156. The views and conclusions contained in this document are those of the authors and should not be interpreted as representing the official policies, either expressed or implied, of the Army Research Office or the U.S. Government. The U.S. Government is authorized to reproduce and distribute reprints for Government purposes notwithstanding any copyright notation herein. L.M.C. acknowledges support of NSF RAPID Grant Number 2029262. S.S. acknowledges support from the Fralin Biomedical Research Institute. S.D. acknowledges a fellowship from the China Scholarship Council (CSC) under Grant CSC Number 201806770029.

## Appendix

### Simulation method

For a fully mixed system, the time evolution of a number of individuals in each state of our modified compartmental SEAIR epidemic model can be prescribed by the following set of deterministic equations:

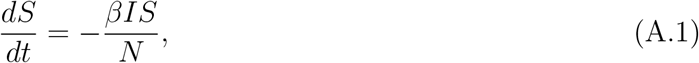

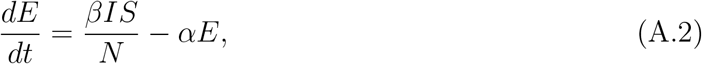

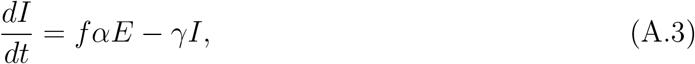

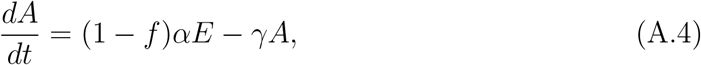

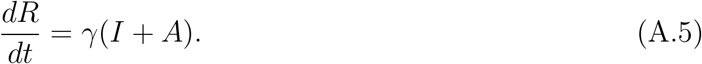

The macroscopic mean-field reaction rates {*β, α, γ*} in these equations are related to the stochastic reaction rates {*r, b, a*} used in the simulations in Fig. 1 in a generally non-trivial manner. Below we show how the simulation reaction rates are approximately related to the mean-field reaction rates. To find the exact numerical factors that enter these relations, a renormalization analysis of the stochastic coarse-grained Langevin equation that describes the epidemic spread on small-world network would be necessary [48].

Microscopically, one can draw the following approximate relations between the stochastic and the mean-field reaction rates:

- A susceptible individual *i* is exposed with a rate ℛ_*i*_ ≈ *rm/n* ≈ *β*×[*fraction of infectious individuals*], where *m* is the number of infectious neighbors (both short and long-range) and *n* is the total number of neighboring sites. Since for parallel sequential updates each individual is being picked only once, the probability that the susceptible individual will become infected in a single Monte Carlo step is = *rm/n*. We set the microscopic value of the infectious rate *r* = 0.6 to obtain ℛ_0_ = 2.5 for our set of simulation parameters.
- An exposed individual becomes infectious symptomatic or asymptomatic with reaction rates *fb* or (1 − *f*)*b*, respectively, which are related to the mean-field reaction rates via *b* ≈ *α*, since both *E* → *I* and *E* → *A* are linear stochastic processes.
- An infectious identified or unidentified individual recovers with the rate *a* ≈ *γ*, since *I* → *R* and *A* → *R* are also linear processes.

The mean-field rate equations fail to capture the effect of fluctuations and spatial and temporal correlations which play a decisive role in the propagation of the infection. To properly account for these intrinsic fluctuations, we implement a stochastic SEAIR epidemic model on a more realistic two-dimensional Newman–Watts small-world network using the following individual-based Monte Carlo algorithm, which also takes into account the unidentified infectious state *A*. All described stochastic rates {*r, b, a*} are implemented as probabilities that a given reaction will occur for the individual of focus, but we refer to them as rates in our description since for parallel sequential updates these reactions are attempted once for each individual per Monte Carlo step:

1. Randomly distribute *N* individuals on the underlying two-dimensional 1000×1000 square lattice with periodic boundary conditions and with a fraction *ϕ* of long-distance links (c.f. Fig. 2), subject to the restriction that each site may only contain at most one individual. A small fraction of the individuals are initially set to be infectious (half of them in the *I* state and the other half in the *A* state), while the remainder of the population is initialized as susceptible to the infection.
2. Perform parallel updates, i.e., sequentially update the state of each site starting from the first one, by maintaining two state vectors to register the previous state **x** and the current state **X** of all sites. The current state **X** is obtained from the previous state **x** by sequentially carrying out the following actions:
  a. For both the dynamic and static setups, if the selected site *x*_*i*_ contains a susceptible *S*, count the number of infectious individuals (both *I* and *A*) among all its short-distance and long-distance neighbors, and update *X*_*i*_ to *E*, i.e., perform *S* → *E* reaction with probability 1 − (1 − *r/n*)^*m*^, where *m* is the number of *S*-*I* and *S*-*A* pairs, and *n* is the total number of neighboring sites. Likewise, in the following, *X*_*i*_ is tacitly updated from the status of **x**.
  b. If the selected site contains an exposed individual *E*, perform the *E* → *I* or *E* → *A* reaction with probabilities *fb* and (1 − *f*)*b*, respectively.
  c. If the selected site contains an infectious individual, perform the *I* → *R* or *A* → *R* reaction with probability *a*.
  d. For the dynamic setups, if the selected site *x*_*i*_ is occupied by any individual after any of the reactions mentioned above have been attempted, a hopping direction is picked randomly among adjacent and long-distance neighbors. After the hopping direction is picked, if a connected lattice site along the hopping direction is empty, i.e., *x*_*j*_ = 0, then the chosen individual is moved to that site with probability *d* = 1. However, if the lattice site along the hopping direction is occupied by any individual, the hopping reaction is not performed.
  e. After one lattice sweep (one Monte Carlo step), **X** is assigned to **x**.
3. After the first ten Monte Carlo steps, begin periodic testing and isolation protocol:
  a. If the number of Monte Carlo steps starting from *t*_0_ = 10 MCS is a multiple of a chosen testing period TP, perform testing of all individuals in the system. Then mark the identified individuals and their neighbors at that moment, and set a delay timer for isolation to DT for the infectious individuals and to DT + DQ for their neighbors.
  b. Check if any individuals have to be isolated, and if yes, place them on quarantine, i.e., prohibit the individual to interact with any other individual in the system, and disable any movement if the system is dynamic for Q Monte Carlo steps.
  c. Check if any individuals have to be released from quarantine, and if yes, enable their ability to interact with other individuals and to move across the system.
4. Repeat the procedures in item 2 and item 3 for a pre-selected total number of Monte Carlo steps.

The origin of the functional form of the probability for a susceptible individual to be infected comes from the fact that for the parallel sequential updates the normalized probability of not catching the infection from an infectious neighbor is (1 − *r/n*). The probability of not getting infected from *m* infectious neighbors is (1 − *r/n*)^*m*^, and the probability of being infected then reads 1 − (1 − *r/n*)^*m*^ ≈ *mr/n*.

We emphasize that the details of the algorithmic implementation (e.g. algorithmic update rules) only quantitatively change the results, but do not qualitatively change the simulation outcomes. For example, we perform an infection reaction only when we pick a susceptible individual. A different simulation algorithm could perform the infection reaction when an infectious individual is picked. Such algorithms lead to equivalent results, assuming an identical ℛ_0_ is chosen, which may necessitate slight adjustments to the rates, *r, a*, and *b*.

## References

[1] Kermack W O, McKendrick A G and Walker G T, 1927 Proceedings of the Royal Society of London. Series A 115 700–721

[2] Brauer F, 2017 Infectious Disease Modelling 2 113–127

[3] Keeling M J and Eames K T, 2005 Journal of The Royal Society Interface 2 295–307

[4] Anderson R and May R 1992, Infectious Diseases of Humans: Dynamics and Control (Oxford University Press, Oxford)

[5] Keeling M and Rohani P 2011, Modeling Infectious Diseases in Humans and Animals (Princeton University Press)

[6] Murray J D 2002, Mathematical Biology, Vols. I + II (Springer, New York, 3rd ed)

[7] Täuber U C 2014, Critical Dynamics – A Field Theory Approach to Equilibrium and Non-Equilibrium Scaling Behavior (Cambridge University Press, Cambridge)

[8] Lindenberg K, Metzler R and Oshanin G (eds) 2019, Chemical Kinetics: Beyond The Textbook (World Scientific Publishing Company)

[9] Newman M E J, Watts D J and Strogatz S H, 2002 Proceedings of the National Academy of Sciences 99 2566–2572

[10] Newman M E J, 2002 Phys. Rev. E 66(1) 016128

[11] Newman M E J and Watts D J, 1999 Phys. Rev. E 60(6) 7332–7342

[12] Eubank S, Eckstrand I, Lewis B, Venkatramanan S, Marathe M and Barrett C L, 2020 Bulletin of Mathematical Biology 82 52–59

[13] Mukhamadiarov R I, Deng S, Serrao S R, Priyanka Nandi R, Yao L H and Täuber U C, 2021 Scientific Reports 11 130–138

[14] Pastor-Satorras R and Vespignani A, 2001 Phys. Rev. Lett. 86(14) 3200–3203

[15] Pastor-Satorras R and Vespignani A, 2001 Phys. Rev. E 63(6) 066117

[16] Salathé M, Kazandjieva M, Lee J W, Levis P, Feldman M W and Jones J H, 2010 Proceedings of the National Academy of Sciences 107 22020–22025

[17] Eames K, Bansal S, Frost S and Riley S, 2015 Epidemics 10 72–77

[18] Nishiura H, Kobayashi T, Miyama T, Suzuki A, Jung S, Hayashi K et al., 2020 International Journal of Infectious Diseases 94 154–155

[19] Tindale L C, Stockdale J E, Coombe M, Garlock E S, Lau W Y V, Saraswat M et al., 2020 eLife 9 e57149

[20] Kraemer M U G, Yang C H, Gutierrez B, Wu C H, Klein B, Pigott D M et al., 2020 Science 368 493–497

[21] Fu H, Xi X, Haowei Wang H, Boonyasiri A, Wang Y, Hinsley W et al. 2020 The COVID-19 epidemic trends and control measures in mainland China Tech. Rep. Imperial College London

[22] Ferguson N M, Laydon D, Nedjati Gilani G, Imai N, Ainslie K, Baguelin M et al. 2020 Impact of non-pharmaceutical interventions (NPIs) to reduce COVID-19 mortality and healthcare demand Tech. Rep. Imperial College London

[23] Delamater P L, Street E J, Leslie T F, Yang Y and Jacobsen K H, 2019 Emerging Infectious Diseases 25 1–4

[24] Hoertel N, Blachier M, Blanco C, Olfson M, Massetti M, Rico M S et al., 2020 Nature Medicine 26 1417–1421

[25] Hunter E, Mac Namee B and Kelleher J, 2018 PLOS ONE 13 1–35

[26] Qin J, You C, Lin Q, Hu T, Yu S and Zhou X H, 2020 Science Advances 6 1–7

[27] He X, Lau E H Y, Wu P, Deng X, Wang J, Hao X et al., 2020 Nature Medicine 26 672–675

[28] McAloon C, Collins Á, Hunt K, Barber A, Byrne A W, Butler F et al., 2020 BMJ Open 10 1–9

[29] Li R, Pei S, Chen B, Song Y, Zhang T, Yang W and Shaman J, 2020 Science 368 489–493

[30] Petersen E, Koopmans M, Go U, Hamer D H, Petrosillo N, Castelli F et al., 2020 The Lancet Infectious Diseases 20 e238–e244

[31] Salje H, Tran Kiem C, Lefrancq N, Courtejoie N, Bosetti P, Paireau J et al., 2020 Science 369 208–211

[32] Li Q, Guan X, Wu P, Wang X, Zhou L, Tong Y et al., 2020 New England Journal of Medicine 382 1199–1207

[33] Mossong J, Hens N, Jit M, Beutels P, Auranen K, Mikolajczyk R et al., 2008 PLOS Medicine 5 1–1

[34] Leung K, Jit M, Lau E H Y and Wu J T, 2017 Scientific Reports 7 7974–7986

[35] Fraser C, Riley S, Anderson R M and Ferguson N M, 2004 Proceedings of the National Academy of Sciences 101 6146–6151

[36] Ódor G, 2021 Phys. Rev. E 103(6) 062112

[37] Kretzschmar M E, Rozhnova G, Bootsma M C J, van Boven M, van de Wijgert J H H M and Bonten M J M, 2020 The Lancet Public Health 5 e452–e459

[38] Grantz K, Lee E, D’Agostino McGowan L, Lee K, Metcalf J, Gurley E and Lessler J, 2021 PLoS Medicine 18

[39] Quilty B J, Cli?ord S, Hellewell J, Russell T W, Kucharski A J, Flasche S et al., 2021 The Lancet Public Health 6 e175–e183

[40] Smith L E, Potts H W W, Amlåt R, Fear N T, Michie S and Rubin G J, 2021 BMJ 372 1–13

[41] He S, Peng Y and Sun K, 2020 Nonlinear Dynamics 101 1667–1680

[42] Mwalili S, Kimathi M, Ojiambo V, Gathungu D and Mbogo R, 2020 BMC Research Notes 13 352–357

[43] Carcione J M, Santos J E, Bagaini C and Ba J, 2020 Frontiers in Public Health 8 230–243

[44] Ames G M, George D B, Hampson C P, Kanarek A R, McBee C D, Lockwood D R et al., 2011 Proceedings of the Royal Society B: Biological Sciences 278 3544–3550

[45] Priyanka and Verma V 2020 (2006.14373)

[46] Bertozzi A L, Franco E, Mohler G, Short M B and Sledge D, 2020 Proceedings of the National Academy of Sciences 117 16732–16738

[47] Dye C, Cheng R C H, Dagpunar J S and Williams B G, 2020 Royal Society Open Science 7 201726

[48] Täuber U C, 2012 Journal of Physics A: Mathematical and Theoretical 45 405002

